# Cost-effectiveness of genetic and clinical predictors for choosing combined psychotherapy and pharmacotherapy in major depression

**DOI:** 10.1101/2020.03.31.20048538

**Authors:** Chiara Fabbri, Siegfried Kasper, Joseph Zohar, Daniel Souery, Stuart Montgomery, Diego Albani, Gianluigi Forloni, Panagiotis Ferentinos, Dan Rujescu, Julien Mendlewicz, Alessandro Serretti, Cathryn M. Lewis

**Affiliations:** Social, Genetic and Developmental Psychiatry Centre, Institute of Psychiatry, Psychology and Neuroscience, King’s College London, United Kingdom; Department of Psychiatry and Psychotherapy, Medical University Vienna, Austria; Department of Psychiatry, Sheba Medical Center, Tel Hashomer, and Sackler School of Medicine, Tel Aviv University, Israel; Laboratoire de Psychologie Medicale, Universitè Libre de Bruxelles and Psy Pluriel, Centre Européen de Psychologie Medicale, Brussels; Imperial College School of Medicine, London, UK; Laboratory of Biology of Neurodegenerative Disorders, Neuroscience Department, Istituto di Ricerche Farmacologiche Mario Negri IRCCS, Milan, Italy; Department of Psychiatry, Athens University Medical School, Athens, Greece; University Clinic for Psychiatry, Psychotherapy and Psychosomatic, Martin-Luther-University Halle-Wittenberg, Germany; Université Libre de Bruxelles; Department of Biomedical and NeuroMotor Sciences, University of Bologna, Italy

**Author notes:** **Corresponding author:** Chiara Fabbri MD PhD, Social, Genetic and Developmental Psychiatry Centre, Institute of Psychiatry, Psychology and Neuroscience, King’s College London, De Crespigny Park, Denmark Hill, London, United Kingdom, SE5 8AF.

**Keywords:** Antidepressants, Genetics, Psychotherapy, Treatment-Resistant Depression, Cost-Effectiveness

## Abstract

**Background:** Predictors of treatment outcome in major depressive disorder (MDD) could contribute to evidence-based therapeutic choices. This study tested the cost-effectiveness of a pharmacogenetic and clinical predictive model (PGx-CL-R) vs a clinical risk (CL-R) predictive model in guiding the assignment of combined pharmacotherapy and psychotherapy vs pharmacotherapy in MDD.

**Methods:** We hypothesized that the prescription of combined treatment, a strategy with evidence of increased efficacy vs pharmacotherapy, may be optimized based on the baseline risk of pharmacotherapy resistance, estimated through PGx-CL-R or CL-R predictive models. Both strategies were compared to standard care (ST, pharmacotherapy to all subjects). Treatment effects, costs and utilities (quality adjusted life years, QALYs) were based on the literature and included in a three-years Markov model.

**Results:** CL-R was cost-effective compared to PGx-CL-R, with ICER (incremental cost effect ratio) of £2341 (CL-R) and £3937 (PGx-CL-R) per QALY compared to ST. PGx-CL-R had similar or better ICER compared to ST only when 1) the cost of genotyping was £100 per subject or less or 2) the sensitivity of the PGx-CL-R test was at least 0.90 and the specificity at least 0.85. CL-R had ICER of £3664 and of £4110 when the CL-R model was tested in two independent samples.

**Limitations:** lack of validation in clinical trial.

**Conclusions:** Prediction of pharmacotherapy resistance according to clinical risk might be a cost-effective strategy if confirmed on large samples from the general population. Combined treatment with drugs having a very good tolerability profile could be a cheaper alternative to psychotherapy.

## 1. Introduction

Major depressive disorder (MDD) is the fourth leading cause of disability worldwide (GBD 2015 Disease and Injury Incidence and Prevalence Collaborators, 2016). Overcoming the trial and error principle in treating MDD has been the object of multiple research efforts, but the heterogeneity of the disorder makes it challenging to identify reliable response predictors (Fried and Nesse, 2015) (Kraus et al., 2019). Pharmacogenetic biomarkers are a potential strategy, but only pharmacokinetic variants are endorsed by guidelines so far (Fabbri et al., 2018), while genetic variants involved in drug pharmacodynamics have an unclear role, probably because of their multiplicity and complex interactions. Pharmacokinetic variants alone explain only a limited part of inter-individual variability in treatment clinical outcomes and genotyping is suggested in those who did not respond to at least one pharmacotherapy, thus potential clinical benefits are limited to a subset of patients who are already or are becoming treatment-resistant (International Society of Psychiatric Genetics, 2019). Another issue is the lack of definitive evidence on the cost-effectiveness of pharmacogenetic testing. Previous studies evaluated the cost-effectiveness of testing individual genetic variants in candidate genes (Olgiati et al., 2012; Perlis et al., 2009; Maciel et al., 2018; Groessl et al., 2018; Hornberger et al., 2015). These studies showed that pharmacogenetic testing may be cost-effective compared to standard care, but results were heterogeneous, mostly because of the different effect size of the considered genetic variant(s) on clinical outcomes to different drugs. Further, none of the previous studies compared pharmacogenetics-guided treatment with treatment guided by clinical risk factors, which is a cheaper option.

Markov models were the most commonly used method to estimate cost-effectiveness in previous studies; they simulate a cohort of patients moving between health states (e.g. depression, remission) over time based on a matrix of transition probabilities that depend from treatment effects (Pierucci and Zarca, 2019). Each health state has a different level of well-being, defined as utility and measured as quality adjusted life years (QALYs), each QALY representing a year lived in perfect health, and different costs associated with it. Utilities and costs are compared between an experimental intervention or treatment strategy and standard care. The experimental treatment strategy is considered as cost-effective vs. standard care when it is more effective at an extra-cost that the society is willing to pay. There is no univocal value defining the willingness to pay threshold, the National Institute for Health and Care Excellence (NICE) provides a threshold of £20,000 per each QALY gained as a very general indication to evaluate cost-effectiveness of new interventions, but other thresholds have been suggested (National Institute for Health and Care Excellence, 2015).

This study aimed to improve our knowledge on the cost-effectiveness of different strategies to guide treatment choice in MDD. We investigated the cost-effectiveness of a pharmacogenetics-clinical risk predictive model (PGx-CL-R) and a clinical risk predictive model (CL-R) vs. standard care in guiding the identification of MDD patients more likely to benefit from combined treatment vs. pharmacotherapy. The predictors used in the PGx-CL-R and CL-R groups were developed to estimate the risk of pharmacotherapy resistance as part of a previous work (Fabbri et al., 2020). Combined treatment was pharmacotherapy combined with psychotherapy, which has evidence of increased efficacy compared to pharmacotherapy only (Cuijpers et al., 2014; Ijaz et al., 2018). Previous studies (Olgiati et al., 2012; Perlis et al., 2009; Maciel et al., 2018; Groessl et al., 2018; Hornberger et al., 2015) estimated the cost/effectiveness of choosing an antidepressant over the other, however there is no convincing evidence supporting the choice of a particular antidepressant based on the genetic profile of a patient (Food and Drug Administration, 2019) and for this reason we chose to use a general risk of pharmacotherapy resistance. Guidelines such as those curated by the NICE recommend to evaluate the prescription of combined treatment in patients who did not respond to at least one previous intervention (NICE, 2018). We hypothesized that patients at risk of pharmacotherapy resistance can be identified at baseline. Our predictive models of pharmacotherapy resistance were developed using machine learning in order to reflect the polygenicity of this trait and improve predictive performance compared to previous cost-effectiveness studies that were based on the results of candidate gene association studies (Fabbri et al., 2020).

## 2. Methods

### 2.1. Predictive models

The predictive models of TRD (treatment-resistant depression) used in this study were developed in a previous study (Fabbri et al., 2020). TRD was defined as lack of response to least two adequate antidepressant drugs during the current depressive episode in patients with MDD according to DSM-IV criteria. The study protocol was approved by the ethical committees of all participating centers and all patients signed an informed consent. Each predictive model was developed in a training set of patients treated with serotonergic antidepressants according to the pharmacology domain reported in the Neuroscience based Nomenclature (Wilson, 2018) (n=269) and the reported ROC curves were obtained by testing the models in an independent testing sample treated with serotonergic antidepressants (n=118) (Supplementary Figure 1). The choice to use models developed in patients treated with serotonergic drugs aimed to reflect the characteristics of patients who would be treated with pharmacotherapy in primary care, having non-severe MDD. This group is expected to benefit more from a baseline prediction of their risk to progress towards TRD, since preventive measures can be implemented.

The clinical risk-guided (CL-R) model included five clinical risk factors independently associated with TRD in the training sample (chronic depression, number of depressive episodes > 3, suicidal ideation, Montgomery and Asberg Depression Rating Scale (MADRS) interest-activity score ≥ 13 and MADRS pessimism score ≥ 7). These two MADRS subscales were determined according to previous studies (Uher et al., 2012). The pharmacogenetic + clinical risk-guided (PGx-CL-R) model included the same clinical risk factors and rare (from whole exome sequencing data) + common genetic variants (from genome-wide data) in 83 genes selected for their effect on TRD in the training set (Supplementary Table 1). Both models were trained using a gradient boosting classifier. Further details are reported in Supplementary Methods. We did not include cytochrome 2D6 and 2C19 genes because variants in these genes are potentially useful to guide drug choice or dose adjustments in patients who did not respond to at least one previous pharmacotherapy (International Society of Psychiatric Genetics, 2019), thus they should not be genotyped at baseline and they provide no guidance on general risk of treatment resistance.

**Table 1:** incremental cost-effectiveness ratio (ICER) for the different treatment strategies tested. Effect difference was measures in QALYs (quality adjusted life years). All results are referred to a 3 years simulation. The reference treatment is standard care in all cases. PGx-CL-R=phamarcogenetics and clinical risk guided group. CL-R=clinical risk guided group.

| Treatment strategy | Sample | Cost difference £ | Effect difference (QALYs) | ICER (£ per QALY) |
| --- | --- | --- | --- | --- |
| PGx-CL-R | Main sample | 1594 | 0.405 | 3937 |
|  | Main sample, sensitivity analysis † | 1353 | 0.266 | 5083 |
| CL-R | Main sample | 941 | 0.401 | 2341 |
|  | Main sample, sensitivity analysis † | 671 | 0.291 | 2311 |
|  | GENDEP | 861 | 0.235 | 3664 |
|  | TRD2 | 864 | 0.210 | 4110 |

† this is the sensitivity analysis performed by changing the threshold for distinguishing TRD and non-TRD patients, see Supplementary Figure 1

### 2.2. Cost and utility estimation

The considered costs included outpatient visits, hospitalizations, liaison mental health service in case of emergency room access and brief psychotherapy (16 sessions) based on data available on the Personal Social Services Research Unit (PSSRU) website for the year 2018 (Personal Social Services Research Unit (PSSRU), 2018a) (Personal Social Services Research Unit (PSSRU), 2018b), while medication costs were based on NHS prices (as of April 2019) (REGIONAL DRUG AND THERAPEUTICS CENTRE, 2019) in GBP. The cost of whole exome sequencing and genome-wide genotyping were based on commercial prices in 2019 in US dollars (Suwinski et al., 2019) and converted in GBP using purchasing power parity (ppp) exchange rates in 2018 (OECD, 2018). Cost units for visits, psychotherapy and days spent in hospital were inflated by 2.1% yearly, based on the mean inflation rate in the United Kingdom (UK) in 2018-2020 (Statista portal, 2019). Indirect costs related to productivity were not included, because they are likely to be captured by the utility weights (QALYs) (Olgiati et al., 2012).

The probability of hospitalization, emergency room access and mean hospitalization duration during a depressive episode were based on hospital episode statistics reported by the NHS for the period 2017-2018 (NHS digital, 2018) and the literature (Citrome et al., 2019). The frequency of visits in each health state was based on the NICE guidelines and the literature (NICE, 2018; Judd et al., 2016; Simon, 2000). The number of psychotherapy sessions was chosen in line with the most part of the previous studies comparing pharmacotherapy vs combined pharmacotherapy and psychotherapy (Cuijpers et al., 2014). An overview of the considered costs is reported in Supplementary Table 2.

Utilities were measured as QALYs. The assigned values were 0.40, 0.67, 0.88 and 0 for the health states of depression, response, remission and death, respectively (Olgiati et al., 2012; Hornberger et al., 2015). A value of 0.88 and not one was considered for remission because of a negative utility associated with treatment (side effects) (Olgiati et al., 2012).

### 2.3. Statistical analysis

A Markov model was created using the R package “heemod” (Pierucci and Zarca, 2019). The time horizon was three years and each cycle lasted 12 weeks which reflects the duration of most clinical trials of antidepressant response in MDD (Jakobsen et al., 2017), including each treatment level of STAR*D which was used to estimate the reduction in response and remission probability after the first cycle (Supplementary Methods). Four health states were considered: depression, response, remission and death by suicide. At the beginning of the simulation, 1,000 depressed patients were included in each strategy arm (Figure 1). After each cycle, transition to another state was regulated by the probabilities described in Supplementary Table 3, which were dependent on the time spent in a certain health state (Figure 2). Transition probabilities for each treatment group (pharmacotherapy and pharmacotherapy combined with psychotherapy) were estimated based on the existing literature, data from the STAR*D trial (Cuijpers et al., 2014; Jakobsen et al., 2017; de Maat et al., 2007; Sinyor et al., 2010) and sensitivity/specificity of the predictive models (Supplementary Methods, Supplementary Tables 3 and 4). Pharmacotherapy included only antidepressants for those who responded or remitted in the first two cycles, while in those who did not remit or respond in the first two cycles the cost of first line pharmacological treatments for TRD according to the Maudsley Prescribing Guidelines in Psychiatry (Taylor et al., 2018) was added. All pharmacological treatments were hypothesized to be continued for the whole duration of the simulation. Transitions were possible between all states, while death was modelled as an absorbing health state (Figure 1). Only death by suicide was considered because other causes of death were expected to have a negligible effect in a time period of three years in an adult population in the UK. We did not consider dropouts because many are expected to re-enter the health care system seeking treatment if their depressive symptoms did not remit and no difference in dropout rates was reported between combined psychotherapy and pharmacotherapy vs. pharmacotherapy (Cuijpers et al., 2014).

**Figure 1:**
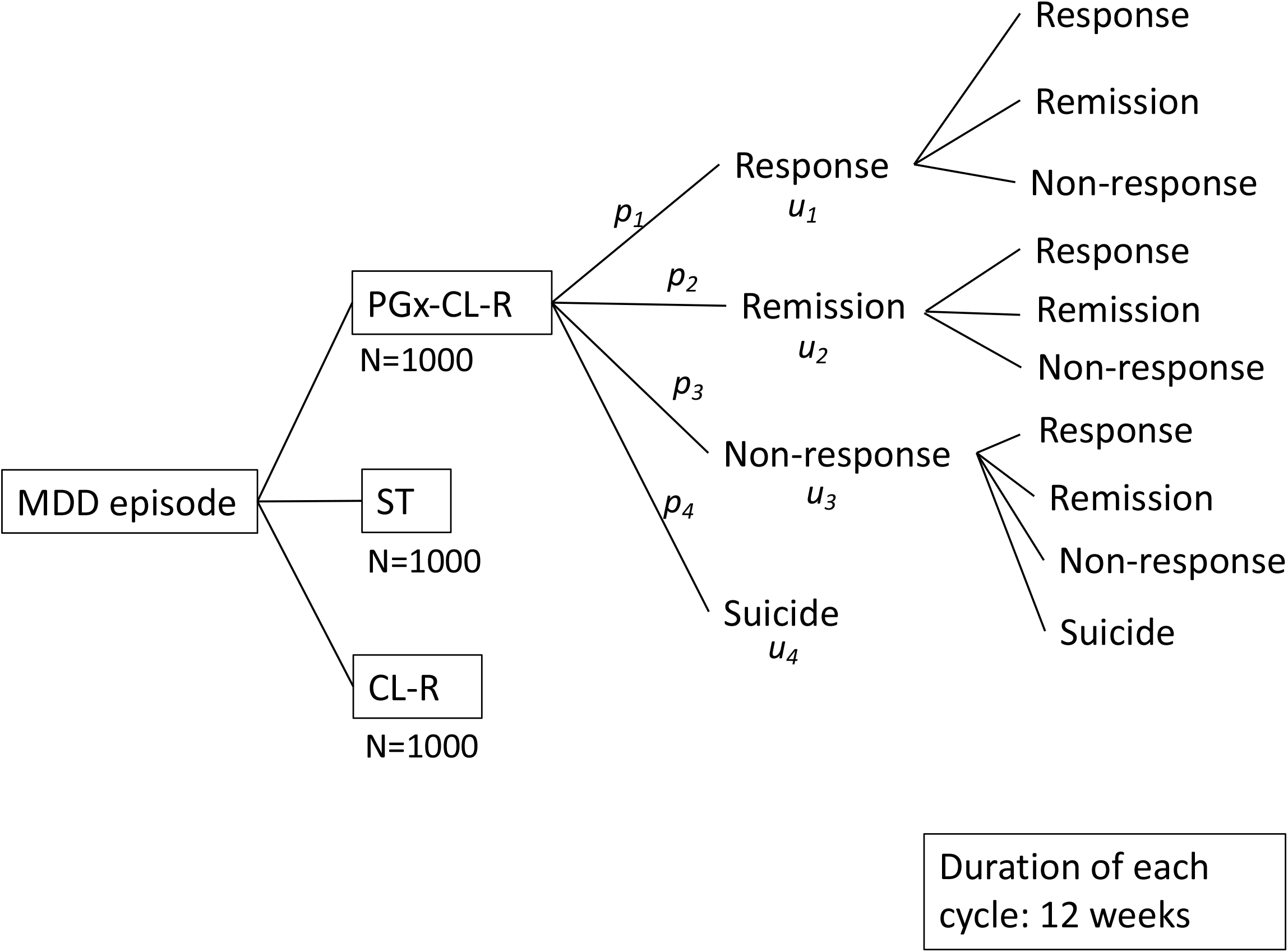
Markov model representation. Each transition is associated with a probability (*p*_*1*_…*p*_*n*_) and each health state with a utility (*u*_*1*_…*u*_*n*_), measured in QALYs.

**Figure 2:**
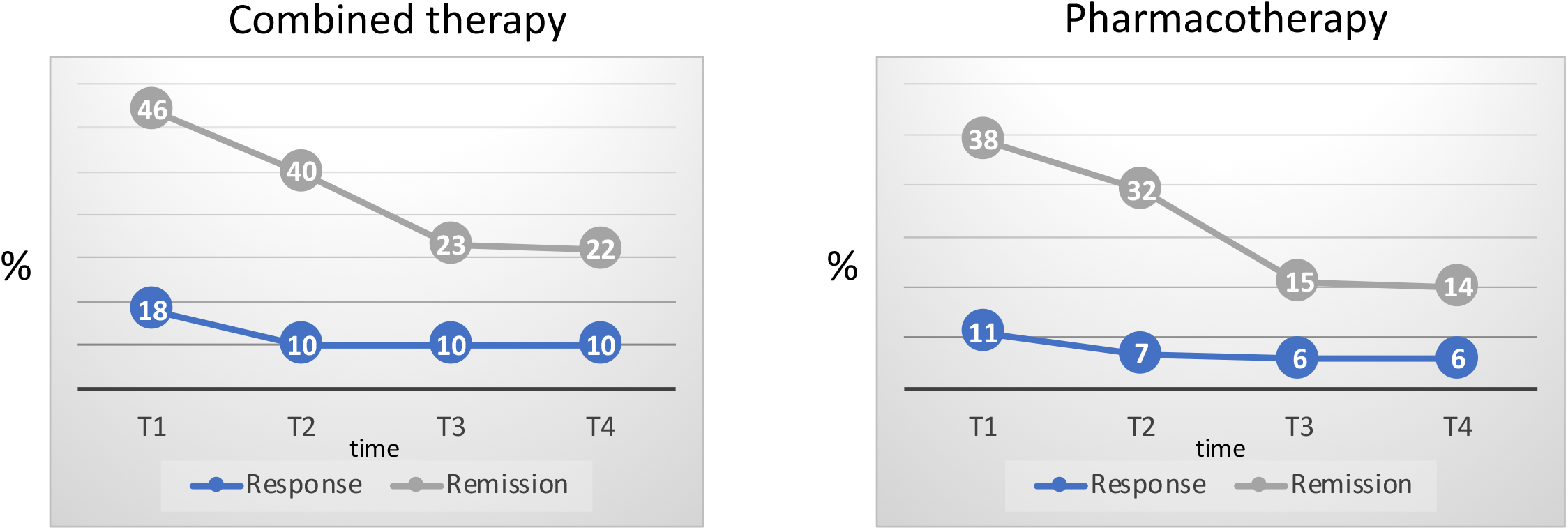
transition probabilities from depression to response and remission in the combined treatment group and pharmacotherapy group. Details on the estimation of these probabilities are reported as Supplementary Methods. T1, T2, T3 and T4: weeks 12, 24, 36, 48.

The two experimental groups were compared to standard care (ST, pharmacotherapy only). In the PGx-CL-R group, subjects at risk of TRD according to the corresponding predictive model were treated with pharmacotherapy combined with 16 sessions of psychotherapy per cycle until response or remission; subjects predicted to be non-TRD were assigned to pharmacotherapy. In the CL-R group, clinical risk factors only were used to assign patients to the combined treatment strategy or pharmacotherapy. For each strategy alternative to ST, we calculated the incremental cost-effectiveness ratio (ICER). ICER is the ratio between the difference in costs and difference in effect (QALYs) of an alternative treatment strategy (*C1* and *E1*) compared to the reference strategy (*C0* and *E0*):

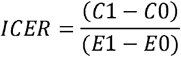

### 2.4. Sensitivity analyses

A probabilistic sensitivity analysis was used to simulate 10,000 cases in each strategy arm for each model parameter (Supplementary Table 5). This generated a probability distribution of ICERs and cost-effectiveness acceptability curves (CEAC) for each treatment strategy.

We also studied the effect of changing key parameters in our model: sensitivity and specificity of the predictive models used in the PGx-CL-R and CL-R groups (Supplementary Figure 1) and costs of sequencing/genotyping. Different combinations of sensitivity and specificity of the PGx-CL-R predictive model were also evaluated to estimate the extent to which hypothetical improvements in the test would change the results.

### 2.5. Replication

The cost-effectiveness of the CL-R strategy was tested in two independent samples, GENDEP and TRD2, but we could not test PGx-CL-R, because of unavailability of whole exome sequence data. Based on sensitivity and specificity of the CL-R predictive model in these samples, we calculated the transition probabilities between the considered health states (Supplementary Table 7). GENDEP was a 12-week partially randomized open-label pharmacogenetic study with two active treatments (nortriptyline and escitalopram). 867 patients with MDD (ICD-10 or DSM-IV criteria) were included and severity of depressive symptoms was assessed at baseline and weekly by scales including the MADRS (Uher et al., 2010). TRD and response were defined according to a previous study (Iniesta et al., 2016). TRD2 included 417 MDD patients (DSM-IV criteria) who had failed to respond to a previous retrospectively assessed antidepressant and were entered into an open two-phase naturalistic trial: in the first phase patients received a 6-week venlafaxine treatment; then those who failed to respond to venlafaxine were treated for a further 6-week period with escitalopram. Depressive symptoms were assessed every two weeks using the MADRS (Souery et al., 2015).

TRD2 and GENDEP studies were approved by the ethical committees of all participating centers and all patients signed an informed consent.

## 3. Results

The clinical-demographic characteristics of the samples are described in Supplementary Table 6. Our analysis suggested that PGx-CL-R is not cost-effective compared to the other treatment strategies, since it showed an ICER of £3937 per QALY when compared to ST, while CL-R had an ICER of £2341 compared to ST (Table 1; number of subjects in each group during the 3 years simulation: Supplementary Figure 2). In our sensitivity analyses, PGx-CL-R would have an ICER comparable to CL-R if: 1) the cost of sequencing/genotyping is £100 or less (ICER would be £2343); or 2) the PGx-CL-R test had sensitivity ≥ 0.90 and specificity ≥ 0.85 (Figure 3). The CEAC showed that if the willingness to pay threshold is £3923 or higher, CL-R has higher chances of being cost-effective compared to the other treatment strategies (Figure 4A). The ICER for CL-R and PGx-CL-R vs. ST were similar when a different threshold was used to distinguish TRD from non-TRD, aiming to maximize specificity (Supplementary Figure 1; Table 1) and we confirmed that CL-R was the most probable cost-effective strategy when the willingness to pay threshold was £3769 (Figure 4B).

**Figure 3:**
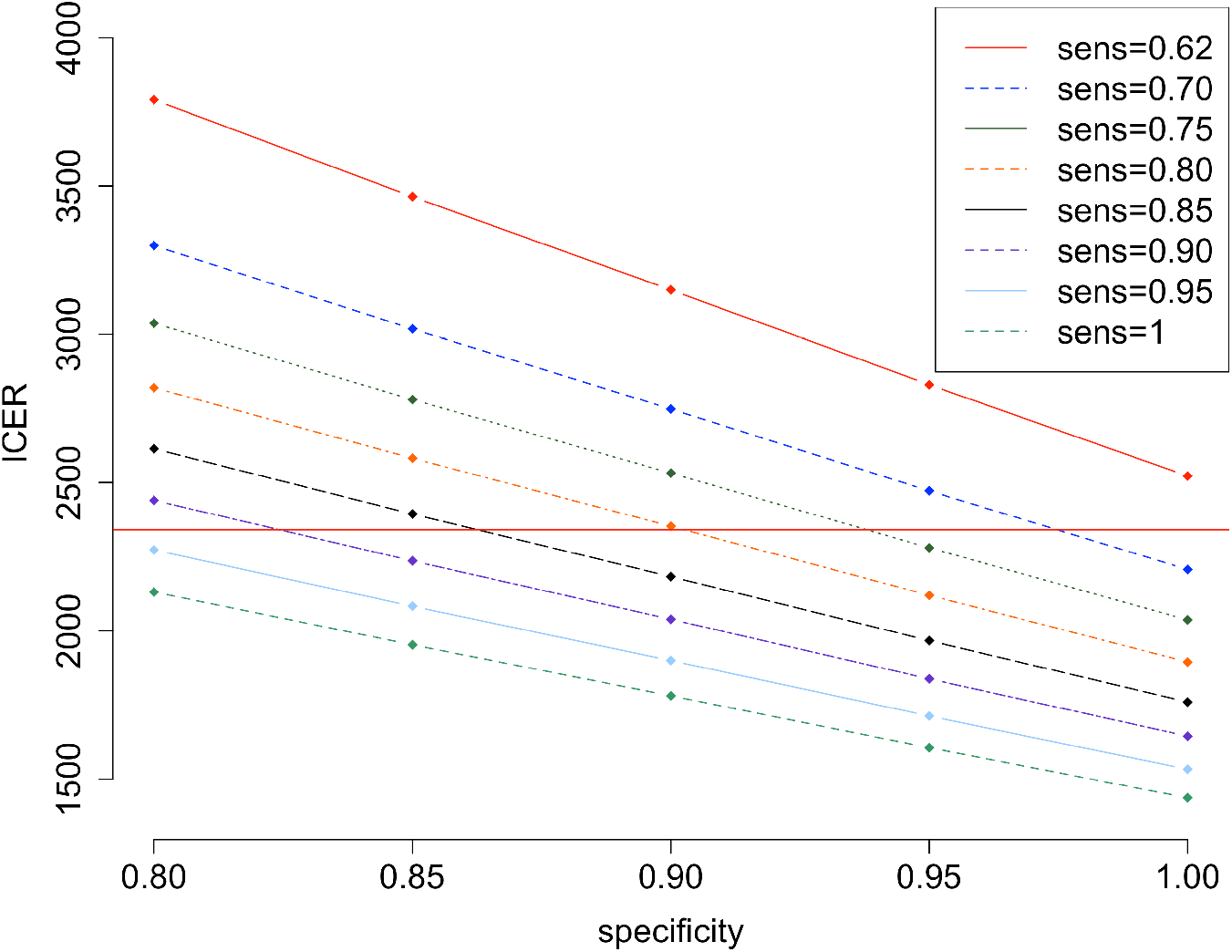
ICER values in the PGx-CL-R group for different combinations of sensitivity and specificity of the predictive test. The ICER of the CL-R group is marked by a red horizontal line for comparison.

**Figure 4:**
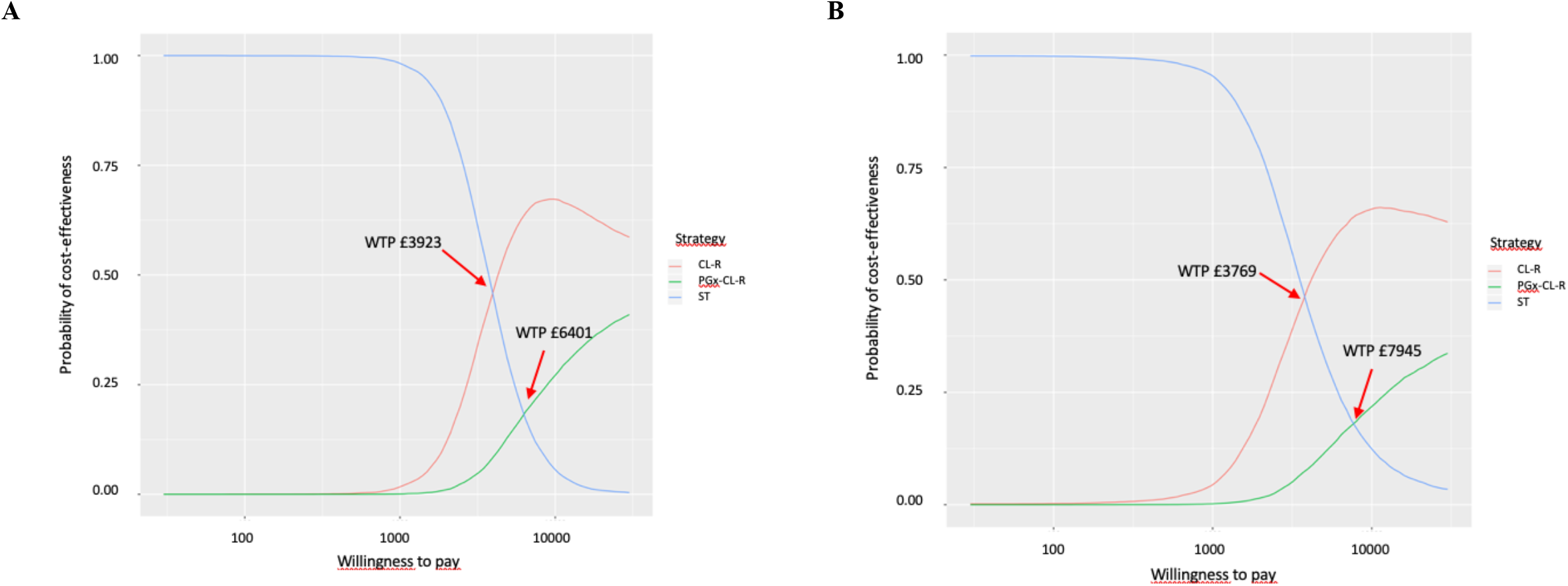
cost-effectiveness acceptability curve (CEAC) for the main analysis (**A**) and in the sensitivity analysis (**B**) maximizing the specificity of the predictive model in the experimental treatment groups (PGx and CL-R). ST=standard care. PGx-CL-R=pharmacogenetics and clinical risk guided group. CL-R=clinical risk guided group. WTP=willingness to pay.

In GENDEP, the CL-R model (Supplementary Figure 1B) had a sensitivity of 0.34 and a specificity of 0.76 and the area under the ROC curve was 0.61 (95% CI 0.55-0.67). The transition probabilities for each treatment strategy are shown in Supplementary Table 7. The ICER of CL-R compared to ST was £3664. According to the CEAC, CL-R had a higher chance of being cost-effective compared to ST when the willingness to pay threshold was £5389 or higher (Supplementary Figure 3A).

In TRD2, the CL-R model (Supplementary Figure 1B) had a sensitivity of 0.30 and a specificity of 0.76 (transition probabilities are in Supplementary Table 7). The area under the ROC curve was 0.56 (95% CI 0.51-0.62). The ICER of CL-R compared to ST was £4110. According to the CEAC, if the willingness to pay threshold was £5942 or higher, CL-R had higher chances to be cost-effective compared to ST (Supplementary Figure 3B).

## 4. Discussion

This study investigated the cost-effectiveness of using a PGx-clinical risk predictive model (PGx-CL-R) vs. a clinical risk predictive model (CL-R) in guiding the identification of MDD patients more likely to benefit from combined treatment vs. pharmacotherapy. Our results suggest that the PGx-CL-R treatment strategy is not cost-effective compared to CL-R, since it provided a specificity only 7-10% higher, at a high cost of exome sequencing and genotyping, which had a major impact on the ICER.

There are two key scenarios which would make the ICER of PGx-CL-R analogous or better than the ICER of CL-R: 1) cost of genotyping/sequencing of £100 or less; or 2) sensitivity of the test of at least 0.90 and specificity of at least 0.85 (Figure 3). It should be noted that commercial pharmacogenetic tests for guiding antidepressant prescription currently cost more than $2000 (Hornberger et al., 2015). A reduction of sequencing/genotyping cost is likely to occur in the next years (National Human Genome Research Institute, 2018) and genotyping could become a standard procedure as part of health care for disease screening and drug response/side effect profiling, and costs would be balanced by the benefits of prevention, early diagnosis and personalized treatments. However, in the near future, clinical risk factors seem the most reasonable strategy to work on.

Clinical variables associated with TRD have been investigated, and a number of severity indicators have been identified (De Carlo et al., 2016). However, few studies have developed predictive models of TRD, and none replicated their predictive performance in independent samples (Iniesta et al., 2016; Kautzky et al., 2018).

We developed a clinical risk score based on only five variables, in order to make assessment as easy as possible, and we included only variables generalizable to most MDD samples. The clinical score showed a fair performance in the testing sample, in which it was estimated to improve the detection of TRD cases of 22% compared to assigning the combined treatment randomly to the same proportion of subjects (62% vs. 40% in 1000 simulations). In the two replication samples the predictive performance was weaker, and the proportion of TRD cases that can be identified was 7% higher (GENDEP) and 5% higher (TRD2) than what would be expected if combined treatment was assigned randomly to the same proportion of subjects, probably as a consequence of different clinical characteristics of the replication samples. It should also be noted that these estimates depend from the prevalence of TRD, which was assumed to be 31% in this study (Supplementary Methods), since the proportion of TRD patients identified by our predictive models compared to chance would improve in samples with lower prevalence of TRD. The reported increase in TRD identification may be seen as a marginal benefit, but the corresponding ICER are good compared to the standard for new interventions (National Institute for Health and Care Excellence, 2015) and MDD is the fourth leading cause of disability worldwide (GBD 2015 Disease and Injury Incidence and Prevalence Collaborators, 2016), justifying the investment of resources in its treatment. Another point in favor of CL-R is that antidepressant augmentation with psychological treatment has a low risk of harming patients’ health.

In the probabilistic sensitivity analyses, we showed that if the willingness to pay threshold is £3800-£5900 (depending on the sample), CL-R is the most probable cost-effective strategy. These values are in a line with the results of a clinical trial that assessed the cost-effectiveness of combined psychotherapy and pharmacotherapy vs. pharmacotherapy in patients with MDD who did not respond to at least one previous pharmacotherapy. This study found that the ICER was £5374 for combined treatment vs. pharmacotherapy after 3.3 years (Wiles et al., 2016). After three years, we showed that similar ICER values would be obtained by assigning treatment group based on a clinical risk score at baseline. However, our results are based on response and costs estimated from the literature and not on observed data from a clinical trial. The lack of real clinical and cost data is the main limitation of this study. Response and remission to combined pharmacotherapy and psychotherapy were based on a meta-analysis of RCTs, which were single-blind in ∼1/3 of cases (the assessors were blind to treatment, but of course the patients were not) (Cuijpers et al., 2014).

A treatment strategy that our study did not explore was the use of pharmacological augmentation to antidepressants instead of augmentation with psychotherapy. However, there is no meta-analysis estimating the effect size of pharmacological augmentation vs. antidepressant only as first line treatment in MDD and in TRD the effect size of this treatment on symptom improvement is similar to that reported for augmentation with psychotherapy (Cuijpers et al., 2014; Ijaz et al., 2018; Zhou et al., 2015; Strawbridge et al., 2019). However, if a drug with very good tolerability profile is demonstrated to have similar or better efficacy than psychotherapy for augmenting treatment in MDD, combined treatment with this drug would be cost-effective compared to combined treatment with psychotherapy.

This study point towards the possibility of using clinical risk factors to guide the prescription of combined psychotherapy and pharmacotherapy after the baseline MDD assessment, instead of waiting to see non-response to the first drug. Given the limited resources available in most clinical settings, the predictive performance should be optimized though studies in large naturalistic cohorts from primary care, in order to identify the best threshold of the clinical risk score in this population.

## Data Availability

This study used previously published results to to create a Markov simulation.

## Role of the Funding Source

Chiara Fabbri is supported by a Marie Skłodowska-Curie Actions Individual Fellowship funded by the European Community (EC Grant agreement number: 793526; project title: Exome Sequencing in stages of Treatment Resistance to Antidepressants - ESTREA).

Cathryn M. Lewis is part-funded by the National Institute for Health Research (NIHR) Biomedical Research Centre at South London and Maudsley NHS Foundation Trust and King’s College London.

The collection of the main sample included in this study and TRD2 sample was supported by an unrestricted grant from Lundbeck for the Group for the Study of Resistant Depression (GSRD). Lundbeck had no further role in the study design, in the collection, analysis, and interpretation of data, in the writing of the report, and in the decision to submit the paper for publication. All authors were actively involved in the design of the study, the analytical method of the study, the selection and review of all scientific content. All authors had full editorial control during the writing of the manuscript and approved it.

The GENDEP project was supported by a European Commission Framework 6 grant (contract reference: LSHB-CT-2003-503428). The Medical Research Council, United Kingdom, and GlaxoSmithKline (G0701420) provided support for genotyping. This paper represents independent research part-funded by the National Institute for Health Research (NIHR) Biomedical Research Centre at South London and Maudsley NHS Foundation Trust and King’s College London. The views expressed are those of the author(s) and not necessarily those of the NHS, the NIHR or the Department of Health and Social Care. High performance computing facilities were funded with capital equipment grants from the GSTT Charity (TR130505) and Maudsley Charity (980).

## Acknowledgements

None

## Potential conflicts of interest

Dr. Souery D. has received grant/research support from GlaxoSmithKline and Lundbeck; has served as a consultant or on advisory boards for AstraZeneca, Bristol-Myers Squibb, Eli Lilly, Janssen and Lundbeck. Prof. Montgomery S. has been a consultant or served on Advisory boards: AstraZeneca, Bristol Myers Squibb, Forest, Johnson & Johnson, Leo, Lundbeck, Medelink, Neurim, Pierre Fabre, Richter. Prof. Kasper S. received grants/research support, consulting fees and/or honoraria within the last three years from Angelini, AOP Orphan Pharmaceuticals AG, Celegne GmbH, Eli Lilly, Janssen-Cilag Pharma GmbH, KRKA-Pharma, Lundbeck A/S, Mundipharma, Neuraxpharm, Pfizer, Sanofi, Schwabe, Servier, Shire, Sumitomo Dainippon Pharma Co. Ltd. and Takeda. Prof. Zohar J. has received grant/research support from Lundbeck, Servier, Brainsway and Pfizer, has served as a consultant or on advisory boards for Servier, Pfizer, Abbott, Lilly, Actelion, AstraZeneca and Roche, and has served on speakers’ bureaus for Lundbeck, Roch, Lilly, Servier, Pfizer and Abbott. Prof. Mendlewicz J. is a member of the Board of the Lundbeck International Neuroscience Foundation and of Advisory Board of Servier. Prof. Serretti A. is or has been consultant/speaker for: Abbott, Abbvie, Angelini, Astra Zeneca, Clinical Data, Boheringer, Bristol Myers Squibb, Eli Lilly, GlaxoSmithKline, Innovapharma, Italfarmaco, Janssen, Lundbeck, Naurex, Pfizer, Polifarma, Sanofi, Servier. Cathryn Lewis is a member of the R&D SAB of Myriad Neuroscience. The other authors declare no conflict of interest.

## Notes

### Clinical Trial

n/a

### Funding Statement

Chiara Fabbri is supported by a Marie Skłodowska-Curie Actions Individual Fellowship funded by the European Community (EC Grant agreement number: 793526; project title: Exome Sequencing in stages of Treatment Resistance to Antidepressants - ESTREA).
Cathryn M. Lewis is part-funded by the National Institute for Health Research (NIHR) Biomedical Research Centre at South London and Maudsley NHS Foundation Trust and King’s College London.
The collection of the main sample included in this study and TRD2 sample was supported by an unrestricted grant from Lundbeck for the Group for the Study of Resistant Depression (GSRD). Lundbeck had no further role in the study design, in the collection, analysis, and interpretation of data, in the writing of the report, and in the decision to submit the paper for publication. All authors were actively involved in the design of the study, the analytical method of the study, the selection and review of all scientific content. All authors had full editorial control during the writing of the manuscript and approved it.
The GENDEP project was supported by a European Commission Framework 6 grant (contract reference: LSHB-CT-2003-503428). The Medical Research Council, United Kingdom, and GlaxoSmithKline (G0701420) provided support for genotyping. This paper represents independent research part-funded by the National Institute for Health Research (NIHR) Biomedical Research Centre at South London and Maudsley NHS Foundation Trust and King’s College London. The views expressed are those of the author(s) and not necessarily those of the NHS, the NIHR or the Department of Health and Social Care. High performance computing facilities were funded with capital equipment grants from the GSTT Charity (TR130505) and Maudsley Charity (980).

